# Evaluating Diuretics in Normal Care (EVIDENCE): Protocol of a cluster randomised controlled equivalence trial of prescribing policy to compare the effectiveness of thiazide-type diuretics in hypertension

**DOI:** 10.1101/2020.12.23.20248767

**Authors:** Amy Rogers, Angela Flynn, Isla S Mackenzie, Lewis McConnachie, Rebecca Barr, Robert WV Flynn, Steve Morant, Thomas M MacDonald, Alexander Doney

## Abstract

**Introduction:** Healthcare systems must use treatments that are effective and safe. Regulators licensed many currently used older medications before introducing the stringent evidential requirements imposed on modern treatments. Also, there has been little encouragement to carry out within-class, head-to-head comparisons of licensed medicines. For commonly prescribed drugs, even small differences in effectiveness or safety could have significant public health implications. However, conventional clinical trials that randomise individual subjects are costly and unwieldy. Such trials are also often criticised as having low external validity. We describe an approach to rapidly generate externally valid evidence of comparative safety and effectiveness using the example of two widely used diuretics for the management of hypertension.

**Methods and Analysis:** The EVIDENCE (Evaluating Diuretics in Normal Care) study has a prospective, cluster-randomised, open-label, blinded end-point design. By randomising prescribing policy in primary care practices, the study compares the safety and effectiveness of commonly used diuretics in treating hypertension. Participating practices are randomised 1:1 to a policy of prescribing either indapamide or bendroflumethiazide when clinically indicated. Suitable patients who are not already taking the policy diuretic are switched accordingly. All patients taking the study medications are written to explaining the rationale for changing the prescribing policy and notifying them they can opt-out of any switch. The prescribing policies’ effectiveness and safety will be compared using rates of major adverse cardiovascular events (hospitalisation with myocardial infarction, heart failure or stroke or cardiovascular death), routinely collected in national healthcare administrative datasets. The study will seek to recruit 250 practices to provide a study population of approximately 50,000 individuals with a mean follow-up time of 2 years. The primary analysis will test for equivalence with a 30% margin in a per-protocol cohort.

**Ethics and Dissemination:** EVIDENCE has been approved by the East of Scotland Research Ethics Service (17/ES/0016, current approved protocol version 4, 28th September 2019). The results will be disseminated widely in peer review journals, guideline committees, National Health Service (NHS) organisations and patient groups.

**Trial registration number:** ISRCTN 46635087; registered pre-results, 11/08/2017.

**Strengths and limitations of this study design:**

- A cluster randomisation design maximises generalisability of results to UK NHS primary care.
- Study interventions with minimal impact on existing NHS workflows should encourage recruitment.
- Development of electronic study search tools and routinely collected data facilitates participation by remote and rural practices.
- One-off policy interventions may have a limited long-term effect on prescribing behaviour.

## Introduction

Formal comparisons of the effectiveness and safety of medicines with similar modes of action and indication are rare[1]. As the number of available medicines increases, high-quality evidence of comparative effectiveness becomes increasingly essential. This problem was the subject of two recent Lancet reviews and accompanying commentary that promoted comparative effectiveness research for public health [2–4].

Randomised placebo-controlled trials (RCTs), often considered the gold standard for generating healthcare evidence, can be cumbersome, expensive, and time-consuming [5]. Such trials often have low external validity because trial participants are likely to differ from patients encountered in usual healthcare practice [6]. As it is traditionally conducted, the randomised controlled trial is not a suitable tool for generating comparative effectiveness evidence at the required scale and speed. A more efficient method that can generate evidence within acceptable boundaries of precision and within reasonable time frames and resources is needed.

The potential for using routinely collected data to generate knowledge within a learning healthcare framework is increasingly recognised [7]. Whilst such data can be used to produce evidence quickly and efficiently, non-interventional research is subject to biases that limit its usefulness for clinical decision making[8]. The result is, despite significant advances in causal inference techniques[9, 10], purely observational research often fails to influence policy and behaviour[8]. Researchers can enhance the value of these large datasets by combining them with the essential features of randomised trials and healthcare system-specific processes.

We describe a hybrid study design that uses routinely collected healthcare data in combination with randomisation to generate high-quality evidence of comparative effectiveness of prescribing policy efficiently and rapidly. Using the example of thiazide-type diuretic medications, widely used for managing hypertension, the EVIDENCE (EValuatIng DiurEtics in Normal CarE) study will compare policies for prescribing indapamide or bendroflumethiazide. Previous work by our group has demonstrated good public support for this type of research[11]. While we have developed the EVIDENCE protocol using the exemplar of diuretic medicines, the infrastructure and methodology will be applicable to the wide range of situations where comparative safety and effectiveness evidence is lacking for treatments commonly used in the NHS.

### Rationale for EVIDENCE

Cardiovascular diseases are the leading cause of death worldwide with high blood pressure being the most common preventable cause, responsible for 54% of strokes and 47% of ischaemic heart disease[12]. Given the high prevalence of hypertension, affecting around 12.5 million people in England in 2015[13], even small differences in the effectiveness and or safety between widely prescribed blood pressure-lowering medications would imply many thousands of potentially avoidable events.

Thiazide and thiazide-like diuretics are a cornerstone of hypertension treatment. The scant evidence for clinically relevant differences between the thiazide and thiazide-like diuretics has been interpreted variably by blood pressure management guidelines internationally[14]. In 2011, the United Kingdom NICE guidelines for managing hypertension[15] stated that “a thiazide-like diuretic, chlortalidone or indapamide should be chosen in preference to a conventional thiazide diuretic such as bendroflumethiazide or hydrochlorothiazide”. The guidelines development group (GDG) conceded that “there were no direct comparisons between the different diuretics with regard to clinical outcomes” and that “the GDG found it difficult to reach firm conclusions regarding the comparative efficacy of different thiazide-type diuretics with regard to blood pressure-lowering” [15]. Before the introduction of this guidance, UK diuretic use for hypertension was almost entirely restricted to bendroflumethiazide[16, 17]. The 2011 guideline was therefore suggesting a significant change to the prevalent prescribing of these drugs. The 2019 guideline update, NG136, still recommends a thiazide-like diuretic in preference to a conventional thiazide[18].

The interpretation of the evidence by the GDG has been disputed[19]. A recent meta-analysis from our group has further supported the lack of evidence in this area[20]. Despite NICE guidance, bendroflumethiazide remains the dominant diuretic prescribed in the UK to treat hypertension[16]. The hypertension community remains in equipoise about diuretic choice, and many local prescribing policies did not take up the NICE recommendation. This situation is likely to have also been influenced by chlortalidone not being readily available in the UK and indapamide being significantly more expensive than bendroflumethiazide[21].

### Objectives

The primary objective of EVIDENCE is to demonstrate if a policy of prescribing bendroflumethiazide for the management of hypertension is equivalent to a policy of prescribing indapamide in terms of both safety and efficacy. Our reason for choosing this equivalence design is that the limited evidence currently available suggests no clinically relevant difference between these two drugs[20]. If no clinically relevant difference is demonstrated, this will justify individual prescribers and healthcare providers choosing which drug to offer based on other relevant factors such as availability, and price.

Our secondary objective is to evaluate the feasibility of the EVIDENCE method for conducting clinical effectiveness research in the NHS. We intend that this methodology be adapted to many disease areas as part of a future learning NHS.

## Methods and analysis

### Study design

EVIDENCE is a cluster-randomised, prospective, parallel-group, blinded outcome study comparing cardiovascular event rates in patients in general practices being prescribed either bendroflumethiazide or indapamide to manage hypertension.

NHS general medical practices using electronic medical records will be invited to take part in the study. The study will recruit practices from a broad range of NHS trust regions, including remote and rural practices that are often excluded from traditional trial participation. The study is designed to be minimally disruptive to practice workflow and uses routine prescribing activities. Recruitment of general practices and randomisation of prescribing policy at the practice level allows the rapid accrual of a large observational study population while incorporating the statistical benefits randomisation. Routinely collected national datasets of hospitalisations and deaths will be used to compare cardiovascular event rates between individuals being treated under each of the two policies. National dispensed prescribing information will be used to facilitate the definition of analysis cohorts. General practice data will be used to gather evidence on potentially important adverse effects such as electrolyte and metabolic disturbances. The study will take place in Scotland where national datasets are well established, although we anticipate that extension to other regions of the United Kingdom would be feasible. Figure 1 shows the EVIDENCE study flow-diagram.

**Figure 1.**
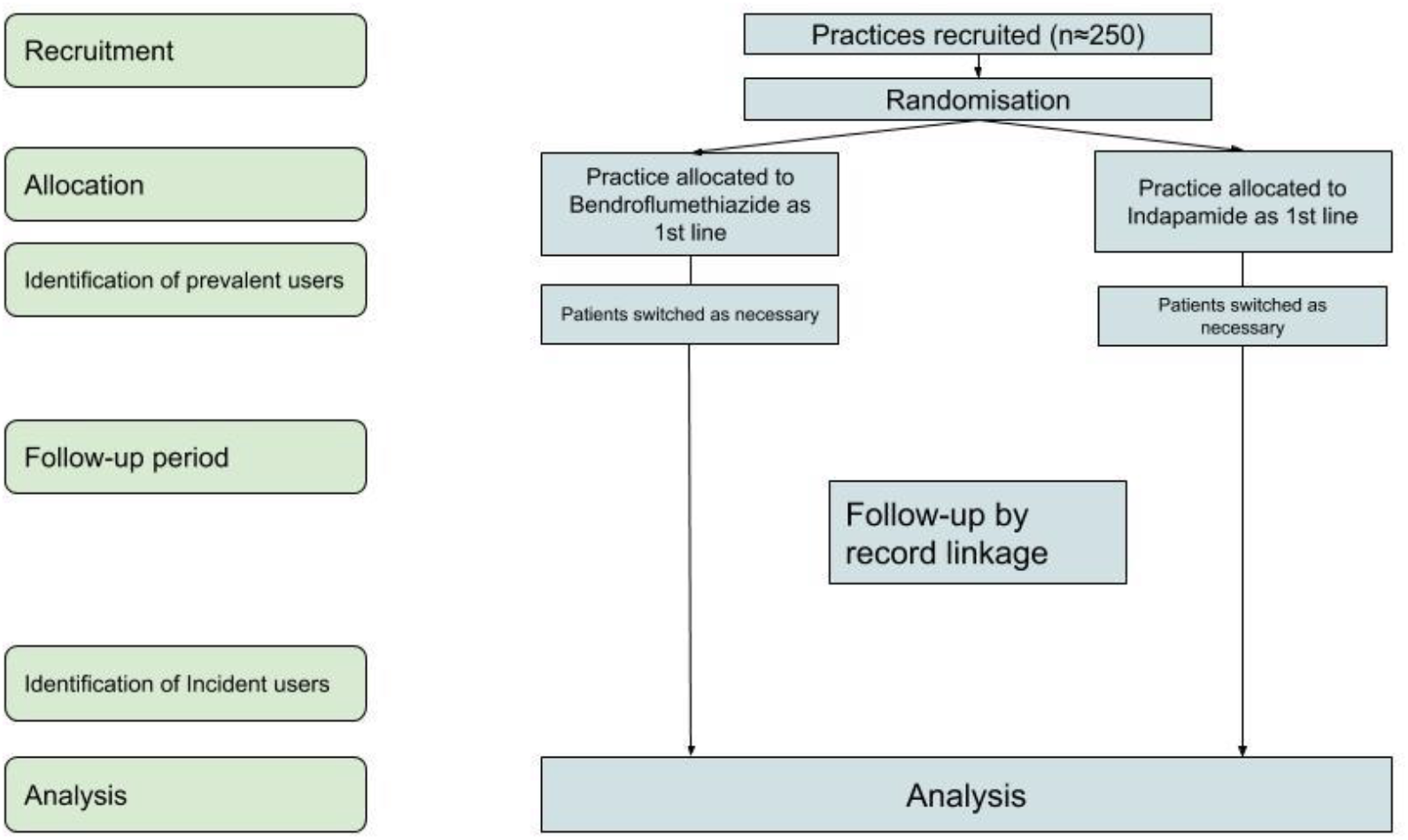
EVIDENCE study flow diagram

### Identification of the study population

A search will be performed in each recruited practice using a bespoke EHR-specific tool to identify patients who may be subject to a medication switch after policy randomisation based on the following criteria.

#### Inclusion criteria

- Documented diagnosis of hypertension (on practice hypertension register)
- Currently receiving repeat prescriptions for bendroflumethiazide or indapamide
- Aged over 18

#### Exclusion criteria

- Documented history of an adverse drug reaction to either medication
- History of having been prescribed both thiazide-like diuretics at different times, indicating a potential clinical reason for not being able to take one or other
- Other cogent clinical or other indication for not switching medication (see below)

A draft list of potential patients identified in each practice will be provided to a relevant member of the primary care team (a practice GP or delegate) for approval. Approval will include the redaction of any individuals considered not suitable for the drug switch, based on local knowledge. The approved list will be used for the study and represents the study population for the practice.

### Cluster Randomisation

The randomisation unit in this study is the practice (the cluster); patients are not individually recruited. Following identification and approval of the practice study population, the practice will be randomised using an online study portal. Randomisation will be 1:1, block balanced by practice list size, using a computerised randomisation algorithm. The prescribing policies will be applied at a practice level, and individual prescribers will remain free to prescribe as clinically appropriate.

### Intervention

#### Policy Medications

- Bendroflumethiazide is licensed for the treatment of hypertension and oedema, although by far the most common indication is hypertension. The typical dose for hypertension is 2.5mg. A small minority of patients are prescribed 5mg or 1.25mg daily.
- Indapamide is licensed to treat hypertension only at 2.5mg/day; although tablets are scored and 1.25mg can be taken. It is also available as a more expensive slow-release preparation at a dose of 1.5mg. The 2.5mg standard release formulation will be used in this study.

### Policy Implementation

#### Informing patients about the study

Immediately following randomisation of a practice, all patients in the study population will be written to informing them that their practice is taking part in the EVIDENCE study. The letter will be printed on practice-specific headed paper and will briefly explain why the study is being conducted. It will also advise that the patient’s repeat prescriptions for thiazide-type medication may be switched in line with the newly assigned practice prescribing policy. The letter directs patients who may have any questions, concerns or objections to visit a study-specific website (www.memoresearch.com/evidence) or directly contact the study team. The study team will provide further information and, if the patient confirms that they would prefer to opt-out of any switch and remain on their current medication, the study team will reverse the switch for that individual.

#### Repeat prescription switching

Patients whose current medication prescription is concordant with the randomly assigned policy will remain on their existing strength and dosage of thiazide-type diuretic. Where a patient is not being prescribed for in concordance with the randomly assigned policy, future repeat prescriptions will be altered as follows:

1. Bendroflumethiazide 2.5mg or 5mg will be changed to indapamide (standard release) 2.5mg.
2. Indapamide 2.5mg will be changed to bendroflumethiazide 2.5mg.
3. Indapamide 1.5mg (slow release) will be changed to bendroflumethiazide 2.5mg.

Practices assigned to Indapamide will also be offered the choice of having any existing Indapamide 1.5mg slow-release prescriptions switched to the more cost-effective 2.5mg version, in keeping with the NICE guidelines.

The study pharmacist/technician will facilitate any prescription changes using established practice prescribing management procedures. All procedures for implementing switches are specified in a study Operations Manual. After the initial switching phase, all patients who newly require a thiazide-type diuretic for hypertension will be subject to the randomly assigned policy. Where possible, integrated electronic practice formularies will be updated to remind prescribers of the policy when issuing new prescriptions for thiazide/thiazide-like diuretics.

### Patient and Public Involvement

During the study pilot, feedback on study materials and methods was sought from patients and healthcare staff who contacted the study team. This feedback was incorporated into improved patient letters and switching methods. Our research unit’s Public Involvement group have discussed the research proposal and design, providing essential guidance on preferred patient communication methods. We will continue to seek Public Involvement group advice as the study is scaled up and when planning dissemination.

### Outcomes

Primary and secondary outcomes will be determined at an individual level and identified by individual-level record linkage of practice study populations to de-identified NHS datasets.

#### Primary Outcome

MACE is a widely accepted composite end-point employed for hypertension trials[22]. This study defines hospitalisation for myocardial infarction, coronary revascularisation, stroke or heart failure, or vascular death.

#### Secondary Outcomes

- Individual components of the primary outcome
- All-cause mortality
- Metabolic complications (hypokalaemia and hyponatraemia)
- New diabetes mellitus diagnoses

#### Tertiary Outcomes

We will evaluate the method in terms of practice workload and acceptability, subsequent prescribing patterns and treatment escalation.

### NHS Data Sets

These data will be provided by the electronic Data Research and Innovation Service (eDRIS), a part of Public Health Scotland. Permission to do this has been obtained from the Public Benefit and Privacy Panel for Health and Social Care (HSC-PBPP). If practices in other parts of the UK are recruited, this information will be obtained through the relevant national agencies.

Dispensed prescribing data for participating practices will be linked to Scottish hospitalisation (SMR01) and National Records of Scotland (NRS) death registration datasets. General practice data will complement these national datasets and allow more accurate ascertainment of secondary outcomes.

All analyses will use anonymised data within a secure research environment, accredited under the NHS Digital - Data Security and Protection Toolkit and the eDRIS National Safe Haven.

### Clinical Adjudication of Cardiovascular Events

The use of routinely collected NHS data for determining cardiovascular end-points for clinical trials is practical and cost-effective. It has been used extensively for large pragmatic trials with careful adjudication of all end-points by a specialised clinical committee[23–26]. However, as administrative data improves, it seems increasingly feasible to use the data without formal adjudication[27, 28]. We intend to adjudicate a proportion of outcome events identified from administrative data to estimate the sensitivity and specificity of administrative diagnostic coding in the study population. This process will require de-anonymisation of the individuals experiencing these events for clinical record retrieval. We have established and standardised procedures to do this based on previous and on-going trials[29].

### Analyses and Statistical methods

The primary analysis will formally test the hypothesis that the two prescribing policies are equivalent. The null hypothesis will be that the two prescribing policies are not equivalent. A primary two-sided equivalence analysis has been chosen over a superiority analysis because a failure to demonstrate statistically significant superiority of one treatment over the other would not be sufficient to prove our objective of equivalence[30]. Figure 2 illustrates the two-sided equivalence analysis.

**Figure 2.**
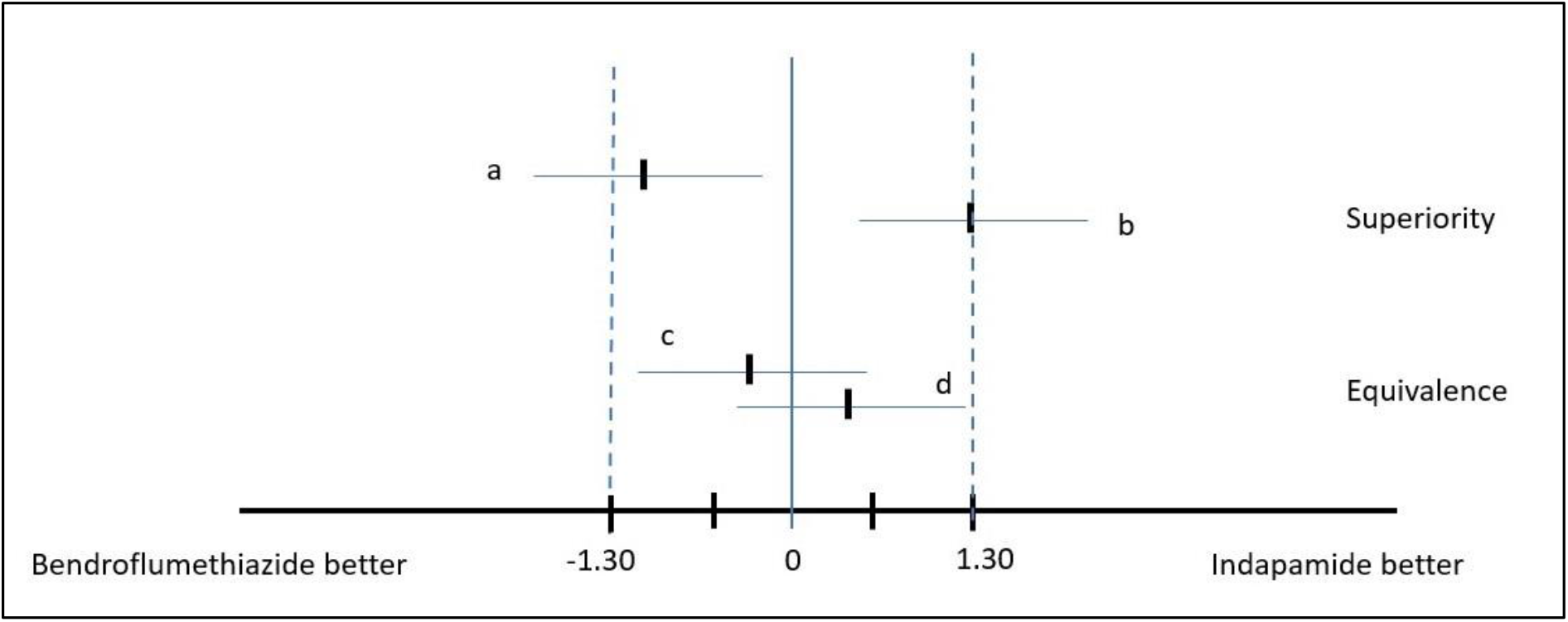
Explanation of equivalence design *(a - bendroflumethiazide superior to indapamide, b - indapamide superior to bendroflumethiazide, c and d - bendroflumethiazide and indapamide are equivalent)*

A systematic review of the effects of thiazide-type and thiazide-like diuretics on cardiovascular events and mortality reported that, in comparison with placebo, thiazide-like diuretics significantly reduced (RR, 0.67 [0.60–0.75]; I2=0%) the number of cardiovascular events[31]. Choosing a relative equivalence margin to preserve ≥50% of this efficacy would dictate an equivalence margin of between 1.25 and 1/1.25. However, it is generally accepted in cardiovascular safety trials to use a margin of between 1.3 and 1/1.3 in line with US Food and Drug Administration (FDA) advice released in 2008 regarding new drugs for Type 2 diabetes[32]. This margin is a reasonable balance between our scientific aims and achievable sample size. Therefore, we selected a 95% confidence interval equivalence margin of 1.3 (i.e., neither medication is either 30% better or 30% worse than the other) based on a two-sided test.

A per-protocol (i.e. on current treatment) analysis is usually preferable in equivalence testing as it is less likely to falsely demonstrate equivalence where protocol adherence is incomplete, despite a real difference in effect; this will be the primary analysis.

We will also perform an intention-to-treat analysis, recognising that results of an analysis reflecting what happened after policy randomisation may be of more direct use to policymakers. We will adjust for a range of pre-specified covariates, including age, sex, co-morbidities, and the Scottish Index of Multiple Deprivation[33].

### Sample Size

As EVIDENCE is a cluster randomisation study, between-practice variation in the end-point is a crucial consideration in determining its sample size[34]. We are not aware of any published methods to account for intra-cluster correlation in a prospective analysis in which follow-up time per patient varies. Therefore, to determine sample size, we modelled data using the Clinical Practice Research Datalink (CPRD). A full description of this modelling is included in supplemental file 1. As part of the Bendroflumethiazide versus Indapamide for Primary Hypertension: Observational (BISON) study[35], we identified patients similar to those we would include in the EVIDENCE study population and found an average 200 individuals per practice. We defined an arbitrary index date and calculated the incidence of the EVIDENCE primary composite end-point (see definition above) following that date. The mean primary outcome event rate was 0.0265 events per patient-year in patients with no history of the outcome, and about six times higher in the 7% of follow-up time in patients with a history. The distribution of event rates between practices was approximately normal, with a standard deviation of 0.0083. However, this variation included within-practice measurement error. We estimate that the variation in underlying event rates between practices had a standard deviation of 0.0047.

We considered plausible relative risks between 1.05 and 1.2 to determine at what level of relative risk the proposed study would be able to declare equivalence with reasonable confidence. We also estimated that recruiting between 100 and 300 practices to this study would be achievable. Combining these assumptions with the BISON study data, we ran simulations that considered relative risks between 1.05 and 1.2, and achievable sample sizes from 100-300 practices. We declared, equivalence if the confidence interval around the point estimate of the relative risk was entirely contained within the interval 1/1.3 - 1.3. We assumed an average follow-up time of 2 years with an anticipated average practice recruitment rate of 5.3 practices per month.

Table 1 summarises the results of the simulation. In a study of 250 practices (study population of ∼50,000) with an average of 2 years follow up per patient, the probability of declaring equivalence between the policies is at least 84% if the point estimate of the relative risk lies between 1/1.15 and 1.15.

**Table 1.**
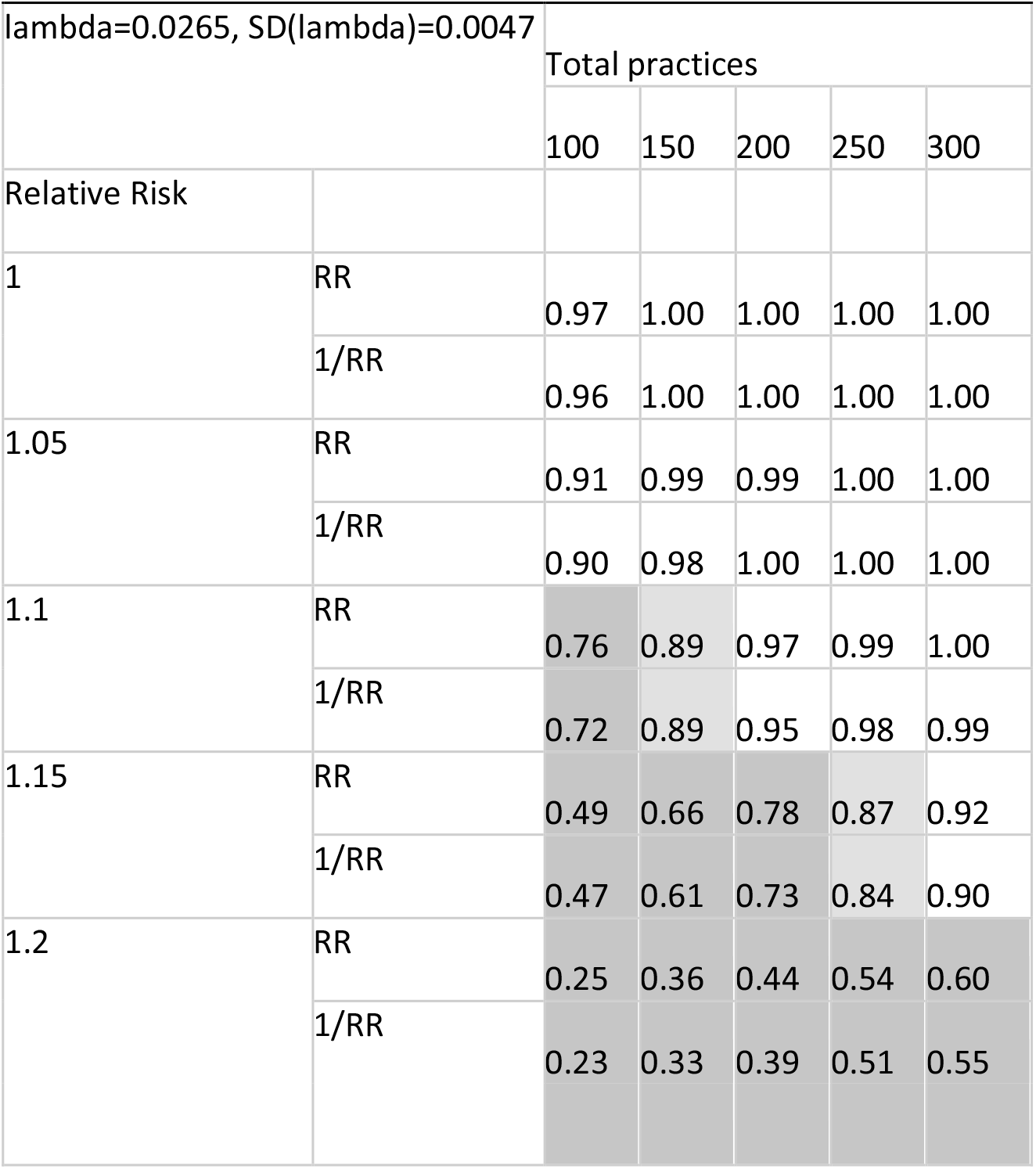
Sample size simulation *(white = >90% probability of declaring equivalence, light grey = >80%, dark grey <80%)*

## Study conduct

### Study Management Group

An executive study committee will be constituted to guide the day to day running of the study. This group will consist of three of the principal investigators. An external advisor will be invited to join the study executive.

### Study Management

A Study Pharmacist and Clinical Research Fellow will oversee the study and will be accountable to the CI. The Study Pharmacist and Clinical Research Fellow will be responsible for checking practice derived data for completeness, plausibility and consistency. However, this remains the overall responsibility of the CI. Any queries will be resolved by the CI or delegated member of the study team. A data monitoring committee will not be constituted as the study compares two prescribing policies, both in common usage.

### Quality Assurance

Since this study intervention is a randomised policy design, and there are no investigational medicinal products, no formal study monitoring is proposed. Principal Investigators and institutions involved in the study will permit quality assurance audits or REC review as required by the Sponsor. In the event of such a review, the Investigators agree to allow the Sponsor, representatives of the Sponsor or regulatory authorities access to all records held by MEMO Research. Where such access is required, persons accessing study data will need to meet the standard conditions permitting such access. It should be noted that all person-specific study data will remain in GP practices and the eDRIS safe haven.

## Ethics and dissemination

The EVIDENCE study will be undertaken by MEMO Research (www.memoresearch.com) and is sponsored by the University of Dundee and NHS Tayside (Sponsor R&D Number 2016CV12). The study has been approved by the East of Scotland Research Ethics Service (REC Number 17/ES/0016) and registered with ISRCTN (46635087). The Medicines and Health products Regulatory Agency has deemed that the EVIDENCE study is not a Clinical Trial of an Investigational Medicinal Product because it is an evaluation of prescribing policies for licensed medications.

### Protocol amendments

Any changes to the study protocol, except those necessary to remove an apparent, immediate hazard, will be reviewed and approved by the chief investigator and sponsor. Amendments to the protocol will be submitted in writing for approval by the appropriate regulatory and ethical authorities before implementation.

### Dissemination

Results of this study will be disseminated through national and international conferences and papers. Authorship criteria will be based on recommendations of the International Committee of Medical Journal Editors. The results will also be shared with guideline committees, NHS organisations and patient groups.

## Discussion

EVIDENCE uses a new pragmatic trial method, combining essential elements of randomised clinical trials and observational analysis of routine clinical data collection as well as making use of routine NHS prescribing management activities. Cluster randomisation allows very large numbers of patients to be rapidly included in a study, facilitating an adequately powered study of short duration. This method provides a research infrastructure for rapid and highly efficient generation of evidence of comparative effectiveness of medications in situations where clinical equipoise exists. Although we have tested the feasibility and acceptability of these methods locally, scaling the study nationally may be challenging. A successful national study will rely on close collaboration between the research team, participating practices, health boards and national data providers. This multi-disciplinary approach to health services research will be vital in achieving a genuinely learning healthcare system.

## Data Availability

No identifiable data will be taken from practices. The data sets used for analysis will be accessible only to approved study staff working through the Public Health Scotland eDRIS safe haven.

## Author’s contributions

The initial idea for this study was conceived by TMM. The current study design was developed by TMM, RF, ISM, AR and AD. The statistical planning and simulations were performed by SM, RF, AD. AR, AF, AD, RB and LM designed and managed the study tools and procedures tested in a feasibility study that has informed to the current protocol. AR, AD and AD drafted the manuscript and all authors have revised and approved the final version of the manuscript.

## Acknowledgements

The authors would like to thank the local prescribing advisors, pharmacists and GP practice staff who contributed their time and expertise to developing study processes. Thanks are also due to the patients and public involvement group members whose feedback has been invaluable.

## Supplemental File

Sample size considerations for the EVIDENCE study

