## Supplemental File 1 for "Evaluating Diuretics in Normal Care (EVIDENCE): Protocol of a cluster randomised controlled equivalence trial of prescribing policy to compare the effectiveness of thiazide-type diuretics in hypertension"

#### Sample size considerations for EVIDENCE study

EVIDENCE is a cluster randomisation study and the between practice variation in the endpoint is an important consideration in determining its sample size.

We extracted data from the CPRD for patients like those we expect to recruit to EVIDENCE. We defined an arbitrary index date and calculated the incidence of the composite endpoint defined for this study following that date, over an average follow up period of 4.7 years per patient. The mean event rate was 0.0265 events per patient year in patients with no history of the outcome, and about 6 times higher in the 7% of follow up time in patients with a history. For patients with no history the distribution of event rates between practices was approximately normal with a standard deviation of 0.0083. However, this variation includes within practice measurement error and we estimate that the variation in underlying event rates between practices had a standard deviation of 0.0047 (see Appendix).

Simulations based on these estimates suggest that in a study of 250 practices with an average of 2 years follow up per patient the probability of declaring equivalence between bendroflumethiazide and indapamide is at least 84% if their relative risk lies between 1/1.15 and 1.15.

### Appendix

The following analyses used 549 CPRD practices with at least 100 patient years of follow up.

Fig 1 shows the distribution of raw event rates in these practices, in patients with no history. It has a mean of 0.0265 events/patient year and a SD of 0.0083.

**Fig 1**

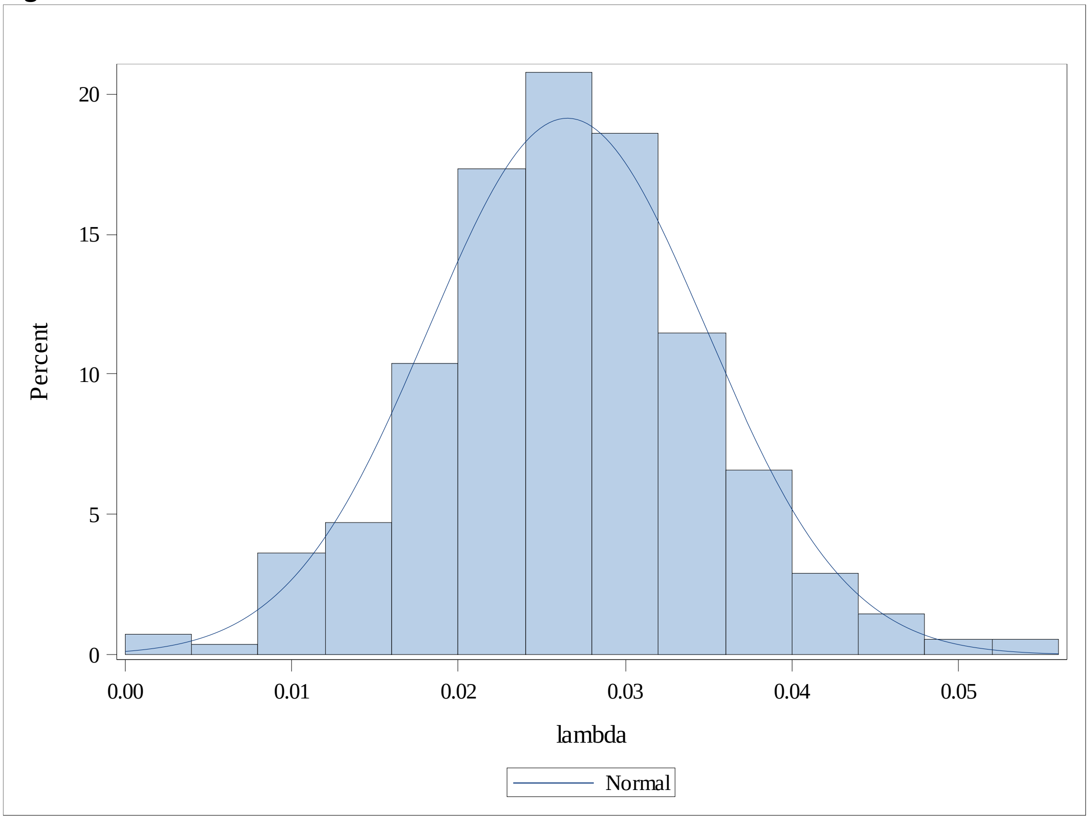


This distribution is the result of both within and between practice variation. Fig 2 shows how the variation decreases with increasing follow up time in the practice. The dots are estimates of the event rate in individual practices. The green line is a 20 point “moving SD“, and the red line is a crudely estimated trend line through these points. The SD approaches an asymptote just below 0.006 (right hand scale) as the practice size increases, although the last 10 or so moving average estimates are lower still at around 0.005. This is a rough estimate of the between practice variation when the within practice variation is small.

**Fig 2**

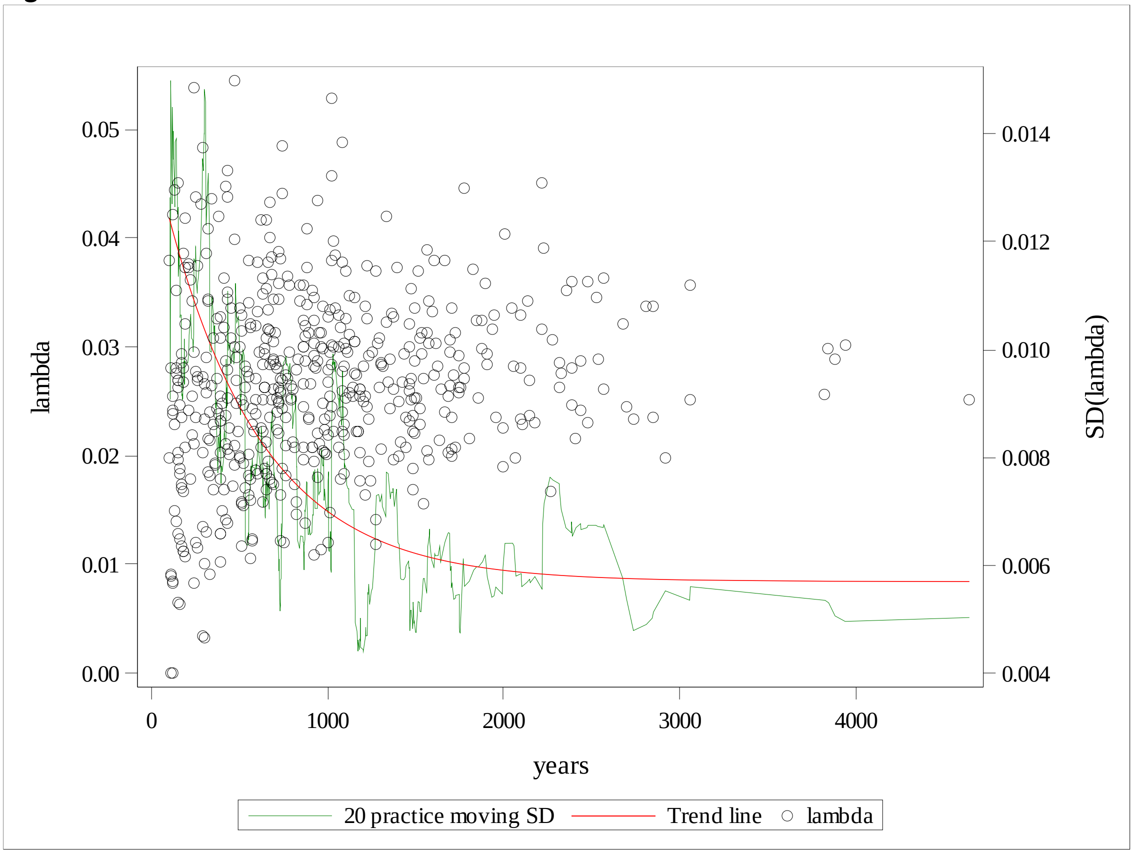


$S_\lambda^2$, the observed variance in event rates between practices, is the sum of the between practice variation in the "true" event rates ($\sigma_B^2$) and the average of the Poisson errors in each estimate ($\sigma_i^2$):

$$S_\lambda^2 = \sigma_B^2 + \frac{1}{n}\sum \sigma_i^2$$

where $n$ is the number of practices.

Since the variance of the number of Poisson events is the number of events, we can estimate the variance of the event rate within each practice as events/years$^2$. The average of these values for the CPRD practices was 0.025923/549 = 0.00004722, and the observed variance between event rates was 0.00006961. The difference is 0.00002239, and the standard deviation for between practice variation in event rates, free from measurement error, is therefore √0.00002239 = 0.0047. This value was used in the simulations.

Using these parameters, we ran simulations to estimate the probability of declaring equivalence in efficacy between bendroflumethiazide and indapamide. For each simulation we selected practices from the CPRD dataset at random, so that variation in practice size was accounted for. We scaled each practice's follow up time by a factor of 2/4.7 to achieve an average of 2 years follow up time per patient. For each practice we generated a random event rate from a normal distribution with mean 0.0265 and standard deviation 0.0047 and used it to generate a random number of Poisson events in 93% of the practice's follow up time. For the remaining 7% of the follow up time we generated events with a 6 times greater rate.

We considered relative risks between 1 and 1.2, and sample sizes from 100 to 300 practices. We generated 1,000 replicates for each combination and for each replicate we used a mixed model to estimate the 95% confidence interval for the relative risk associated with one drug vs the other. We declared equivalence if this confidence interval was contained entirely within the interval 1/1.3 to 1.3. The results are summarised in Fig 3 and Table 1.

**Fig 3 Probability of declaring equivalence vs number of practices**

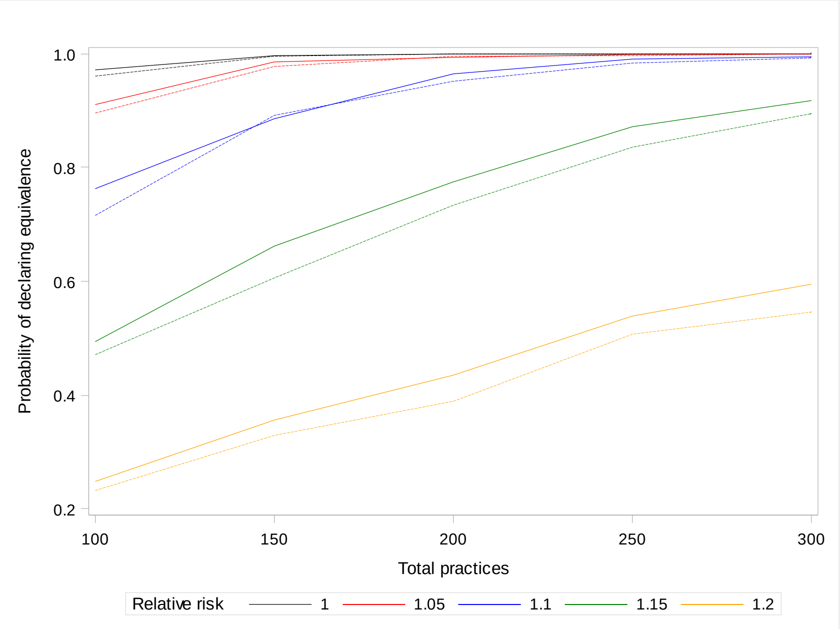


Dotted lines are for 1/RR, i.e. a reduced event rate vs the reference rate of 2.65%/yr.

| **Tables 1** | | | | | | |
|---|---|---|---|---|---|---|
| **lambda=0.0265, SD(lambda)=0.0047** | | **Total practices** | | | | |
| | | **100** | **150** | **200** | **250** | **300** |
| **Relative Risk** | | | | | | |
| **1** | **RR** | 0.97 | 1.00 | 1.00 | 1.00 | 1.00 |
| | **1/RR** | 0.96 | 1.00 | 1.00 | 1.00 | 1.00 |
| **1.05** | **RR** | 0.91 | 0.99 | 0.99 | 1.00 | 1.00 |
| | **1/RR** | 0.90 | 0.98 | 1.00 | 1.00 | 1.00 |
| **1.1** | **RR** | 0.76 | 0.89 | 0.97 | 0.99 | 1.00 |
| | **1/RR** | 0.72 | 0.89 | 0.95 | 0.98 | 0.99 |
| **1.15** | **RR** | 0.49 | 0.66 | 0.78 | 0.87 | 0.92 |
| | **1/RR** | 0.47 | 0.61 | 0.73 | 0.84 | 0.90 |
| **1.2** | **RR** | 0.25 | 0.36 | 0.44 | 0.54 | 0.60 |
| | **1/RR** | 0.23 | 0.33 | 0.39 | 0.51 | 0.55 |

In a study with 250 practices the probability of declaring equivalence between bendroflumethiazide and indapamide is 87% if their true relative risk is 1.15 and 84% if it is 1/1.15.
